# Trends in National Opioid Prescribing for Dental Procedures among Medicaid Patients

**DOI:** 10.1101/2021.04.12.21255158

**Authors:** Ilya Okunev, Julie Frantsve-Hawley, Eric Tranby

**Author notes:** Corresponding Author: Ilya Okunev, 465 Medford Street, Charlestown, MA 02129.

## Abstract

**Background:** We examine trends in opioid prescriptions by dentists for Medicaid-enrolled children and non-senior adults.

**Methods:** We utilized the IBM Watson Medicaid claims databases for 2012-2019 and the CDC conversion dataset. Opioid prescriptions were linked to a dental visit when prescribed within three days of the dental visit and if the patient had no medical visit reported during that period. We conducted descriptive analyses for age, procedures performed, treatment history, and prescription strength.

**Results:** Our study showed consistent decreases in opioid prescription rates in dentistry during the study period: from 2.7% to 1.6% among children (0-20), and from 28.6% to 12.2% for adults (21-64). The adult opioid prescription rate fell for nonsurgical dental procedures from 9.7% to 2.9%. For surgical procedures, the adult prescription rate fell from 48.0% to 28.7%. Most dental-related opioids were prescribed for oral surgeries (Children: 70.8%, Adults: 58.6%). By 2019, 23% of all opioid prescriptions for children were dental related.

**Conclusion:** We found that opioid prescription rates in dentistry for Medicaid enrollees declined significantly between 2012 and 2019 for both children and adults. The percent of prescriptions written for nonsurgical visits consistently declined over the observed timeframe. At the same time, opioid prescription rates for both dental surgical procedures and dental nonsurgical procedures decreased as well.

**Practical Implications:** Although the trends revealed in our analysis show declining opioid prescription patterns, these results suggest that the overall rate is still too high and prescriptions are being written unnecessarily.

## Introduction

The use of opioid medications to control both acute and chronic pain is well established. There were 255.2 million opioid prescriptions written in the United States (US) in 2012, an annual rate of 81.3 prescriptions per 100 persons, according to the Centers for Disease Control and Prevention (CDC)^1^. This fell to 153.3 million or 46.7 prescriptions per 100 persons in 2019.^1^ Although efficacious when used and monitored appropriately, when misused, opioid medications are a leading cause of morbidity, mortality, and have significant social and economic costs. Opioids, both prescribed and illicit, were the leading cause of drug overdose deaths in 2017, according to the CDC, and were involved in 47,800, or 68 percent, of all overdose associated deaths.^2^ For 2017 and 2018 inclusive, the federal government allocated almost $11 billion to address the epidemic of opioid abuse.^3^ From 1999 to 2013, state Medicaid programs spent more than $72.4 billion to combat the epidemic, $8 billion in 2013 alone.^4^ The total economic burden of the opioid epidemic between 2018 and 2020 was reported to be at least $631 billion by the Society of Actuaries.^5^

Negative health outcomes, abuse-related comorbidities, and escalating medical complications and costs are associated with the misuse of opioid medications. Dentists are among the top prescribers of opioid medications and the top prescribers for patients age 10 to 19.^6^ Meanwhile, patients receiving oral health care in an emergency department were more than seven times more likely to receive an opioid prescription than were patients treated in a dental office.^7^ Among all dental patients, surgical, implant and root canal procedures were associated with the highest rates of opioid prescriptions.^8^ Among adolescents and young adults, dental-related visits are often the first exposure to opioid medications.^9-11^ One study revealed that 11.2 % of dentist-prescribed opioids were written for patients younger than 21.^11^ When exposed in high school, young adults are 33% more likely to suffer from future opioid misuse.^12^ More than half of all dental prescriptions remain unused three weeks following a dental surgical extraction and may lead to non-medical misuse.^13^ It is estimated that up to 1,500 deaths annually are due to therapeutically provided dentist-derived prescriptions.^14^

CDC guidelines were released in 2016 for chronic pain management, recommending no more than 10 mg of hydrocodone/acetaminophen tablets every 6 hours for three days. The maximum quantity converted to MMEs translates to no more than 120 MMEs total per prescription.^15^ A 2020 study based on over half a million adult dental visits found that 29.3% of dental opioid prescriptions exceeded the MME-based recommendation, while 53% exceeded the recommended days’ supply.^16^

Hydrocodone, an opium-based analgesic, is associated with the greatest number of opioid deaths.^17^ Together with oxycodone, it remains one of the most commonly prescribed medications by dentists.^18^ Hydrocodone, oxycodone, codeine, and tramadol are associated with high incidence of opioid-related adverse effects.^19^ Tramadol was associated with higher rates of hospitalization, a need for escalated care, and death than other medications, particularly among adolescents.^19^ Due to the associated medical complications of both codeine and tramadol, the Food and Drug Administration on April 20, 2017, issued guidance recommending no use of either medication in children younger than twelve.^20^

Given the stark social and economic costs and the higher likelihood of opioid use disorder and mortality, it is imperative to understand the impact and role of dentists and associated prescribing trends within the child and the non-elderly adult populations, especially for those receiving Medicaid, the insurance program that covers those among the most vulnerable in society. Over 10% of dental visits for those enrolled in Medicaid were associated with an opioid prescription in one study.^21^ Among Medicaid enrollees with opioid use disorder, the percentage without a current opioid prescription has increased from 32% in 2005 to 38% in 2015, despite targeted efforts and programs designed specifically for this population.^22^ In Montana, 28% of unintentional opioid deaths among adults were Medicaid members.^23^ While many studies have evaluated the overall impact of opioids, prescribing practices, and even particular diagnostic subsets, including dental services, there are not many studies that review only Medicaid patients, and fewer still evaluate specific trends and patterns in that population at the national level.

In the present study, we evaluate trends in opioid prescriptions associated with a dental procedure, along with demographic and patient characteristics of patients in a nationally representative Medicaid population. Rates of prescriptions within the population are calculated. Additionally, we review specific prescription types, dosages, morphine milligram equivalents (MMEs), and specific procedures associated with opioids.

## Methods

### Data Source and Sample Selection

For this retrospective analysis, we used deidentified medical, dental, and pharmaceutical claims data from January 1, 2012 through December 31, 2019 from the IBM Watson MarketScan Multi-State Medicaid Database core data set.^24^ The database contains all individual claims from 10 million dental patients among 13 states enrolled in Medicaid across all six years and is considered nationally representative of the overall US Medicaid population. This research was determined exempt from review by the Western Institutional Review Board.

All patients aged 0 to 64 with at least one dental claim between January 1, 2012 and December 31, 2019 were selected. Patients 65 and older and those enrolled in both Medicare and Medicaid (dual-eligibles) were excluded from this analysis, as claims paid by Medicare are not included within this dataset. All opioid medications were included in the analysis with the exception of 2 categories: morphine sulfate and meperidine HCL solutions were excluded due to their use as anesthetic agents.^25^ Buprenorphine/naloxone, buprenorphine, methadone hydrochloride, buprenorphine hydrochloride, morphine sulfate/naltrexone hydrochloride, naloxone hydrochloride/pentazocine hydrochloride were removed because they treat opioid dependence.^26^

### Measures

We employed a methodology previously used in a similar study in a commercially insured population from 2010 through 2015, authored by Gupta and colleagues.^27^ We defined an opioid prescription as dental-related if the prescription occurred either on the same day as a dental visit, or within three days prior to or after the visit. However, if a medical inpatient or outpatient visit happened within the same three days of the prescription, we excluded it as the reason for the prescription could not be determined as dental-related (see Figure 1). We then calculated the percentage of patients receiving an opioid prescription in a calendar year by dividing the number of dental patients receiving at least one opioid prescription by the total number of patients receiving a dental service.

**Figure 1:**
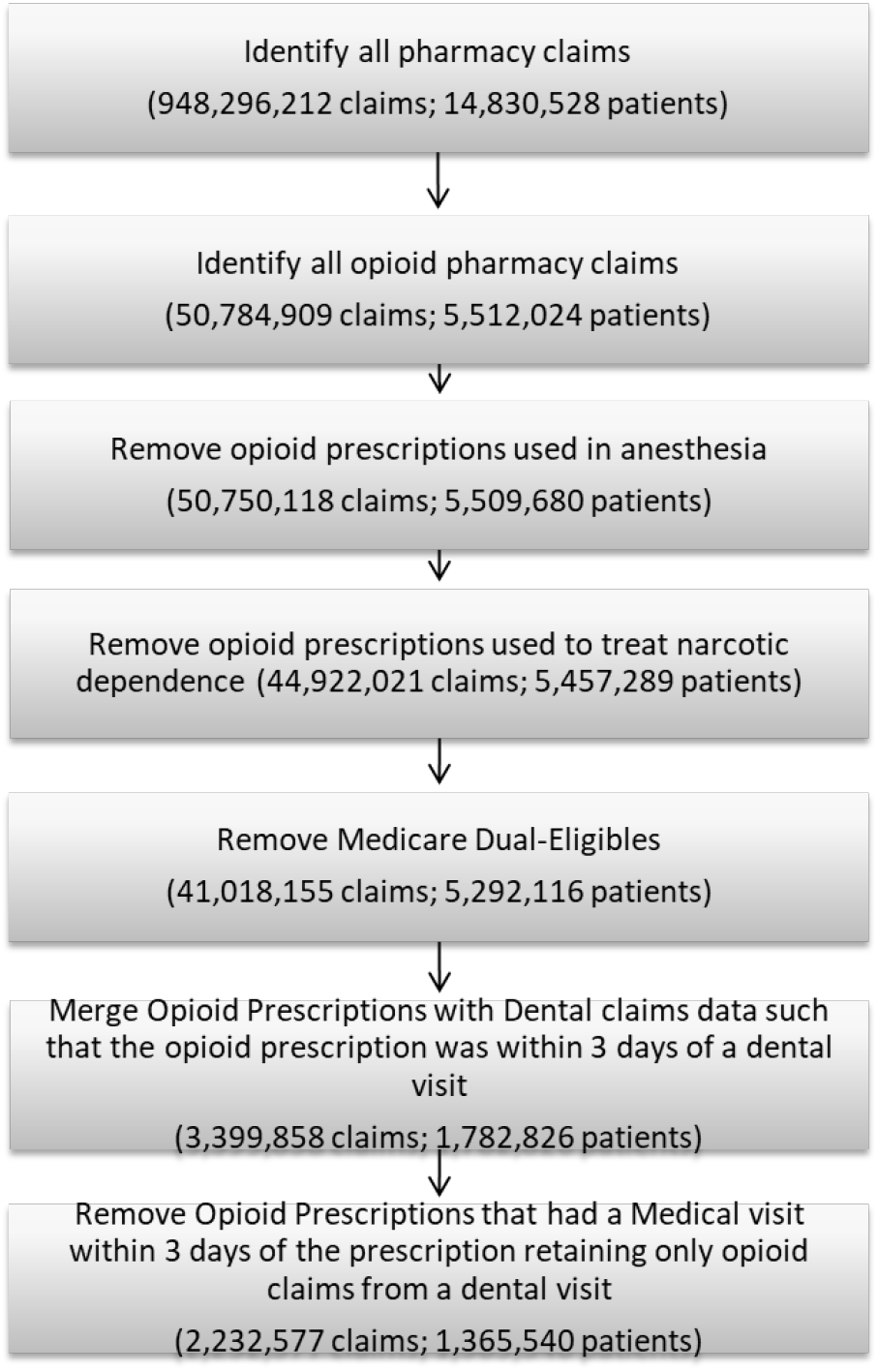
Selection Criteria for Opioid

Due to differences in coverage, eligibility criteria and guidance on prescribing of dental opioids, we stratified patients into two age groups: child patients aged 0 to 20 and adult patients aged 21 to 64. Children and young adults under 21 who are enrolled in Medicaid have mandated dental coverage and are thus eligible for Early and Periodic Screening, Diagnostic, and Treatment benefits (EPSDT), along with other covered screenings and health services. However, dental coverage is optional under federal law for adults (21+) and varies substantially by state.

To estimate the total available opioid morphine milligram equivalency (MME) doses per day, we normalized all prescriptions using a CDC conversion table.^28^ The normalization process allows comparison across opioid medications as the formulations vary considerably in strength. Aligned with CDC recommendations, MMEs were classified into four groups: 0-19.9 MMEs, 20-49.9 MMEs, 50-89.9 MMEs, and 90 or greater MMEs.

As there is a strong association between surgical dental visits and the receipt of opioids, we categorized dental visits as surgical or nonsurgical. Surgical visits were determined by the presence of a surgical procedure code, as defined by past research.^27^ If a surgical code was not present in the claim record, we classified the visit as nonsurgical.

We calculated the rate of prescriptions per service category using the American Dental Association’s CDT categories. Restorative procedures were codes from D2000 to D2999, endodontic (D3000-D3999), periodontic (D4000-D4999), prosthodontic (D5000-D6999), and oral and maxillofacial surgery (D7000-D7999). We combined the CDT categories for implants, fixed prosthodontics, and removable prosthodontics into the general prosthodontics category.

All analysis was performed using SAS 9.4.

## Results

The number of opioid prescriptions per 100 patients per year decreased each year between 2012 and 2019 for both children and adults (Table 1). Among adults enrolled in Medicaid, we saw an overall decline of 68%, from 46.4 prescriptions per 100 patients in 2012 to 15.0 prescriptions per 100 patients in 2019. Among children, we saw an overall decline of 48%, from 3.3 prescriptions per 100 patients in 2012 to 1.7 prescriptions per 100 patients in 2019. Similar reductions were also seen in the percentage of patients receiving an opioid in a given year, declining among adults from 28.6% (CI: 28.5% to 28.8%) in 2012 to 12.2% (CI: 12.1% to 12.2%) in 2019 and from 2.7% (CI: 2.7% to 2.7%) to 1.6% (CI: 1.6% to 1.7%) in children. The median daily MMEs for all age groups was 27.5 for all opioid dental prescriptions between 2012 and 2019. The proportion of prescriptions above 50 MMEs per day declined from 15.5% in 2012 to 3.4% in 2019 with the biggest drops taking place in 2018 and 2019 (Figure 2).

**Table 1:** Dental Opioid Prescriptions.

|  | 2012 | 2013 | 2014 | 2015 | 2016 | 2017 | 2018 | 2019 | Total |
| --- | --- | --- | --- | --- | --- | --- | --- | --- | --- |
| **Child (0-20)** |  |  |  |  |  |  |  |  |  |
| Dental Patients | 2,775,570 | 2,848,933 | 2,895,115 | 3,021,006 | 3,045,902 | 3,065,100 | 2,526,247 | 2,516,665 | 22,694,538 |
| Opioid Prescriptions | 92,410 | 88,335 | 83,809 | 84,655 | 80,527 | 73,058 | 48,977 | 43,821 | 595,592 |
| Dental Patients that received an opioid rx | 75,283 | 73,766 | 72,234 | 74,692 | 72,233 | 66,900 | 45,533 | 41,245 | 521,886 |
| Percentage of patients who received opioid Rx for a dental visit | 2.71%<br>(2.69% to 2.73%) | 2.59%<br>(2.57% to 2.61%) | 2.49%<br>(2.48% to 2.51%) | 2.47%<br>(2.45% to 2.49%) | 2.37%<br>(2.35% to 2.39%) | 2.18%<br>(2.17% to 2.20%) | 1.80%<br>(1.79% to 1.82%) | 1.64%<br>(1.62% to 1.65%) | 2.30%<br>(2.29% to 2.31%) |
| Prescriptions per 100 Dental Patients | 3.33 | 3.10 | 2.89 | 2.80 | 2.64 | 2.38 | 1.94 | 1.74 | 2.62 |
| **Adult (21-64)** |  |  |  |  |  |  |  |  |  |
| Dental Patients | 460,912 | 477,777 | 602,474 | 713,780 | 726,533 | 733,899 | 764,059 | 736,736 | 5,216,170 |
| Opioid Prescriptions | 213,710 | 210,762 | 246,721 | 269,533 | 242,536 | 203,295 | 139,567 | 110,861 | 1,636,985 |
| Dental Patients that received an opioid rx | 131,992 | 132,681 | 161,495 | 184,290 | 173,399 | 151,624 | 109,683 | 89,528 | 1,134,692 |
| Percentage of patients who received opioid Rx for a dental visit | 28.63%<br>(28.50% to 28.77%) | 27.77%<br>(27.64% to 27.89%) | 26.80%<br>(26.69% to 26.91%) | 25.82%<br>(25.71% to 25.92%) | 23.86%<br>(23.77% to 23.96%) | 20.66%<br>(20.57% to 20.75%) | 14.35%<br>(14.28% to 14.43%) | 12.15%<br>(12.08% to 12.23%) | 21.75%<br>(21.72% to 21.79%) |
| Prescriptions per 100 Dental Patients | 46.4 | 44.1 | 41.0 | 37.8 | 33.4 | 27.7 | 18.3 | 15.0 | 31.4 |
| **Total (0-64)** |  |  |  |  |  |  |  |  |  |
| Dental Patients | 3,236,482 | 3,326,710 | 3,497,589 | 3,734,786 | 3,772,435 | 3,798,999 | 3,290,306 | 3,253,401 | 27,910,708 |
| Opioid Prescriptions | 306,120 | 299,097 | 330,530 | 354,188 | 323,063 | 276,353 | 188,544 | 154,682 | 2,232,577 |
| Dental Patients that received an opioid rx | 207,275 | 206,447 | 233,729 | 258,982 | 245,632 | 218,524 | 155,216 | 130,773 | 1,656,578 |
| Percentage of patients who received opioid Rx for a dental visit | 6.40%<br>(6.38% to 6.43%) | 6.20%<br>(6.18% to 6.23%) | 6.68%<br>(6.66% to 6.71%) | 6.93%<br>(6.91% to 6.96%) | 6.51%<br>(6.49% to 6.54%) | 5.75%<br>(5.73% to 5.78%) | 4.72%<br>(4.69% to 4.74%) | 4.02%<br>(4.00% to 4.04%) | 5.93%<br>(5.93% to 5.94%) |
| Prescriptions per 100 Dental Patients | 9.46 | 8.99 | 9.45 | 9.48 | 8.56 | 7.27 | 5.73 | 4.75 | 8.00 |

**Figure 2:**
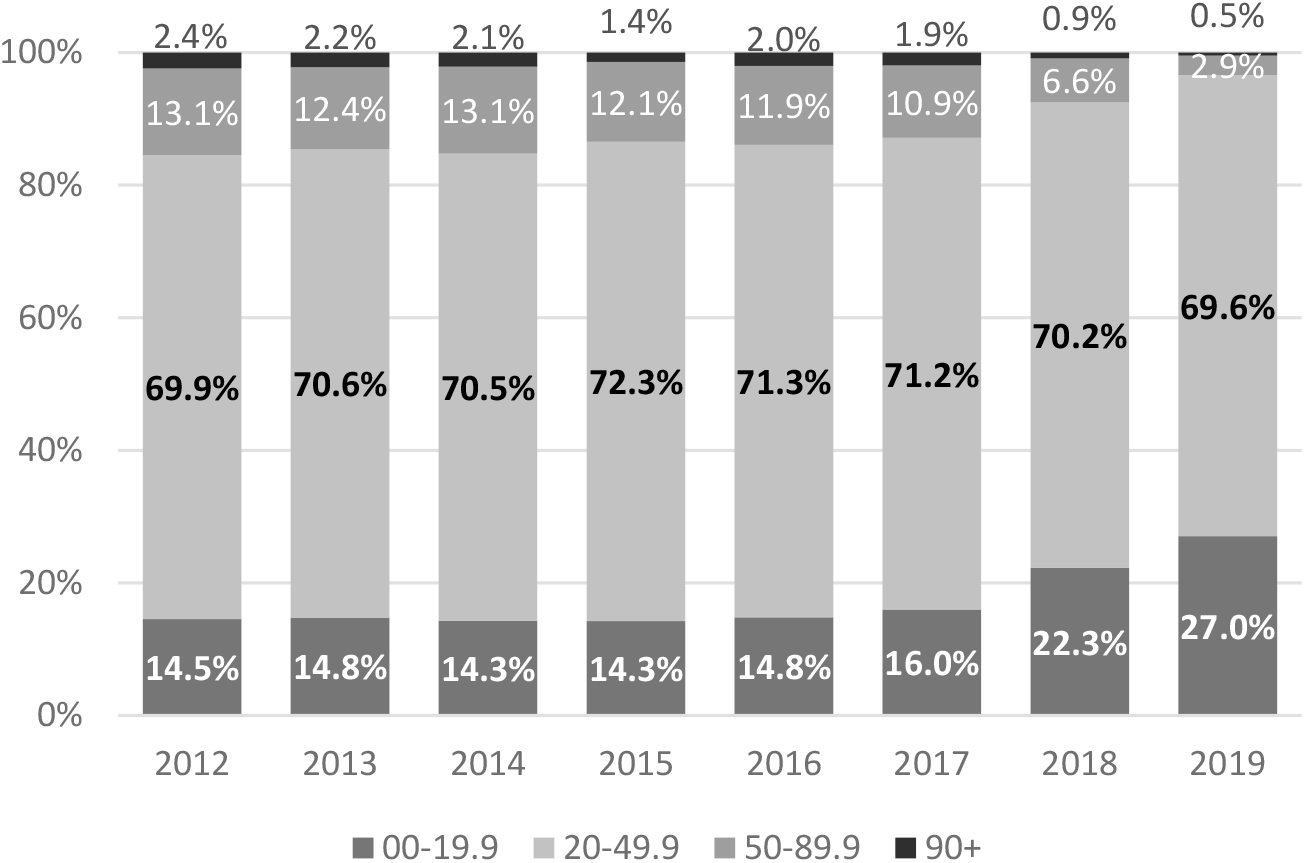
MME/ Day Categories by Year

In our sample of Medicaid patients, 5.4% of total opioid prescriptions were associated with a dental visit between 2012 and 2019. For children, the proportion of opioid prescriptions that are dental related increased from 13.1% in 2012 to 23.0% in 2019. For adults, dental related opioid prescriptions represent a smaller share of total opioid prescriptions with only 4.3% being dental related in 2019 (see Figure 3).

**Figure 3:**
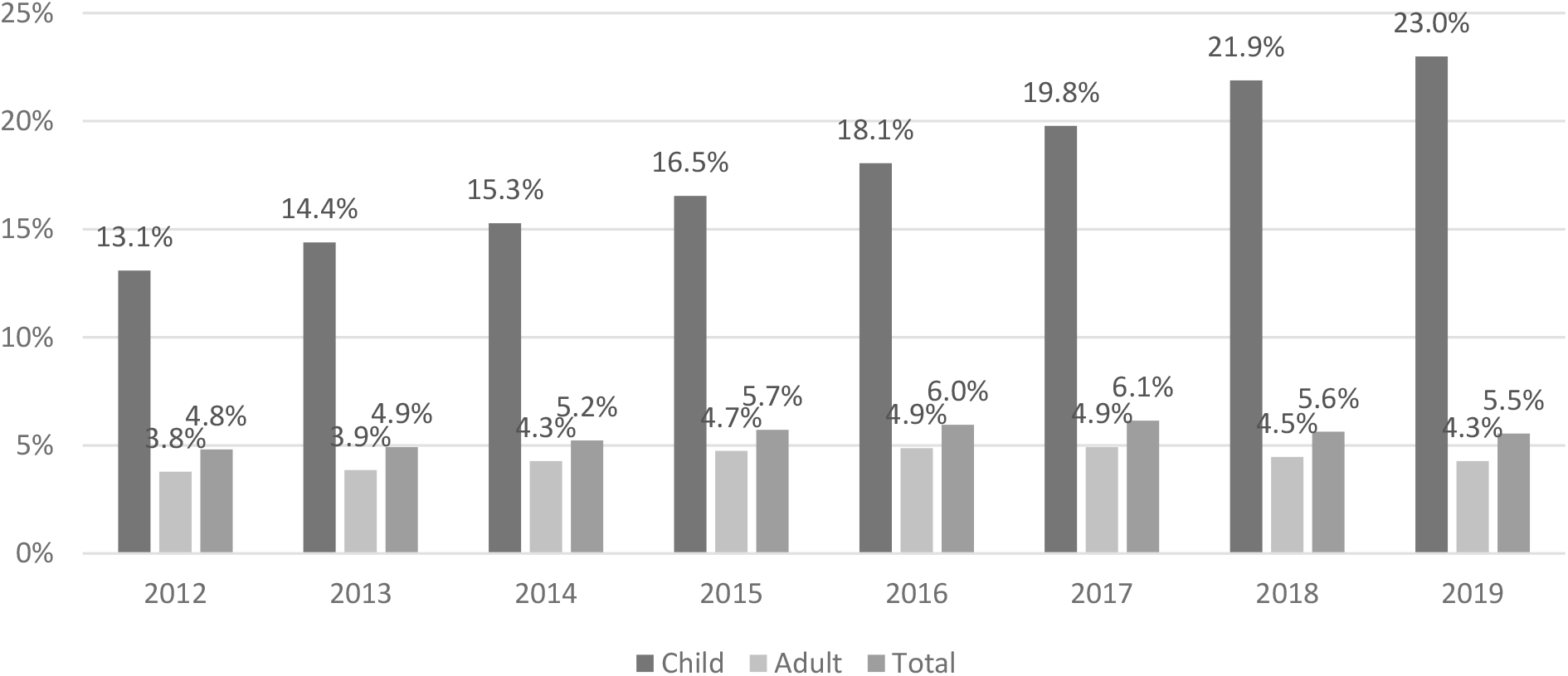
Percent of Opioid Rxs that are Dental Related

Based on CDC’s MME-based guidelines for chronic pain (33), 25.5% of adult opioid prescriptions exceeded the recommended limits for acute pain of 120 total MMEs. The percent of prescriptions over 120 MMEs declined from 33.1% in 2012 to 16.4% in 2019. When considering CDC’s recommendations based on documented days’ supply, the percent of prescriptions for adults over 3 days’ duration dropped from 45.0% in 2012 to 41.6% in 2019 (See Appendix A). It is worth noting that these CDC recommendations are made for chronic pain and are not specifically meant to serve as guidelines for acute dental-related pain.

Mixed results were seen when stratifying by age, race, and gender. Among children, females generally had higher prescription rates than males, with white females having the highest prescription rates overall across all six years in our study. Among adults, white males had the highest overall prescription rates. However, black females had higher prescription rates than white females. Hispanics consistently had the lowest prescription rates overall across all years, ages, and genders analyzed (see Table 2).

Most opioid prescriptions were associated with oral surgery, followed by endodontic and restorative procedures. 76.0% of child opioid prescriptions were given for oral surgery while 63.2% of adult opioids were prescribed for oral surgery. Endodontic procedures accounted for 5.8% of all child dental-related opioid prescriptions and 6.7% of adult prescriptions. Restorations accounted for only 3.3% of child opioid prescriptions and 4.9% of adult prescriptions (see Table 3).

**Table 3.** Percent of Opioid Prescriptions Prescribed for Each Procedure Category.

|  |  |  |
| --- | --- | --- |
| **PROCEDURE CATEGORY** | **Number** | **%** |
| **Children (0-20)** |  |  |
| Oral Surgery | 449,497 | 76.0% |
| Endodontic | 34,183 | 5.8% |
| Restorative | 19,753 | 3.3% |
| Adjunctive General | 10,390 | 1.8% |
| Periodontic | 1,508 | 0.3% |
| Orthodontics | 1,503 | 0.3% |
| Prosthodontics | 156 | 0.03% |
| Total | 591,261 |  |
| **Adults (21-64)** |  |  |
| Oral Surgery | 1,025,604 | 63.2% |
| Endodontic | 109,087 | 6.7% |
| Restorative | 79,894 | 4.9% |
| Adjunctive General | 17,267 | 1.1% |
| Periodontic | 13,722 | 0.8% |
| Prosthodontics (Removable) | 12,207 | 0.8% |
| Prosthodontics (Fixed) | 155 | 0.01% |
| Orthodontics | 137 | 0.01% |
| Total | 1,623,321 |  |
| Each category is counted one time per visit. When a surgical visit occurred, nonsurgical procedures were excluded. Diagnostic and preventive categories were excluded from this table. |  |  |

The prescription rate for nonsurgical encounters declined significantly from 9.7% (CI: 9.6% to 9.7%) in 2012 to 2.9% (CI: 2.9% to 3.0%) in 2019 for all adult encounters across the same time period. Specific to adult surgical encounters, the opioid prescription rate fell from 48.0% (CI: 47.8% to 48.2%) to 28.7% (CI: 28.5% to 28.9%) in the same timeframe. (Table 4).

**Table 4:** Opioid Rx Rate for NonSurgical and Surgical Encounters by Age Group.

|  |  |  |  |
| --- | --- | --- | --- |
| **YEAR** | **Child** | **Adult** | **Total** |
| **NonSurgical Encounters** |  |  |  |
| **2012** | 0.56% (0.55% to 0.57%) | 9.68% (9.61% to 9.74%) | 1.79% (1.78% to 1.80%) |
| **2013** | 0.50% (0.49% to 0.50%) | 9.20% (9.14% to 9.26%) | 1.67% (1.66% to 1.68%) |
| **2014** | 0.45% (0.44% to 0.45%) | 8.74% (8.68% to 8.79%) | 1.77% (1.76% to 1.78%) |
| **2015** | 0.40% (0.40% to 0.41%) | 7.79% (7.75% to 7.84%) | 1.73% (1.72% to 1.74%) |
| **2016** | 0.36% (0.36% to 0.37%) | 6.79% (6.74% to 6.83%) | 1.53% (1.52% to 1.54%) |
| **2017** | 0.28% (0.28% to 0.28%) | 5.43% (5.39% to 5.47%) | 1.22% (1.22% to 1.23%) |
| **2018** | 0.19% (0.19% to 0.19%) | 3.73% (3.70% to 3.77%) | 0.88% (0.87% to 0.89%) |
| **2019** | 0.14% (0.14% to 0.14%) | 2.94% (2.91% to 2.97%) | 0.67% (0.66% to 0.68%) |
| **Surgical Encounters** |  |  |  |
| **2012** | 12.50% (12.41% to 12.59%) | 48.03% (47.84% to 48.23%) | 24.07% (23.97% to 24.17%) |
| **2013** | 12.53% (12.44% to 12.62%) | 47.82% (47.63% to 48.02%) | 24.23% (24.13% to 24.32%) |
| **2014** | 12.45% (12.35% to 12.54%) | 45.29% (45.12% to 45.46%) | 25.32% (25.23% to 25.42%) |
| **2015** | 12.58% (12.49% to 12.67%) | 45.27% (45.11% to 45.43%) | 26.31% (26.21% to 26.40%) |
| **2016** | 12.33% (12.24% to 12.42%) | 43.16% (43.00% to 43.32%) | 25.18% (25.09% to 25.27%) |
| **2017** | 11.90% (11.81% to 11.99%) | 38.95% (38.79% to 39.11%) | 22.99% (22.90% to 23.07%) |
| **2018** | 10.21% (10.12% to 10.31%) | 32.68% (32.52% to 32.85%) | 19.60% (19.50% to 19.69%) |
| **2019** | 9.61% (9.52% to 9.70%) | 28.69% (28.52% to 28.86%) | 17.33% (17.24% to 17.42%) |

Codeine prescriptions in children under 12 declined from 0.55 prescriptions per 100 patients in 2012 to 0.03 prescriptions per 100 patients in 2019. This downward trend preceded FDA guidelines that came out in April of 2017 recommending against prescribing Codeine or Tramadol for pain management in children under 12. Furthermore in 2017, Codeine prescription rates fell by almost 50% from 0.16 prescriptions per 100 patients in Q1 to 0.09 prescriptions per 100 patients in Q4. Tramadol prescription quantities were insignificant for children under 12 for all years between 2012 and 2019 (See Appendix D).

## Discussion

Results of this study show Medicaid-enrolled patients have steadily declining opioid prescription rates since 2012, particularly among adults. Given the increased likelihood of misuse and abuse for prescriptions when written for above 50 MMEs per day, our results are encouraging and show a decrease from 15.5% in 2012 to 3.4% as a percent of total in 2019 in prescriptions written for that level. Also encouraging, these results represent a decline in those opioids prescribed for nonsurgical visits from a 9.7% prescription rate for adults in 2012 to 2.9% in 2019. While our findings demonstrate progress is being made, they also suggest that more improvements are necessary to reduce the rate even further through targeting of particular prescription categories.

Our analysis shows that in 2019, 23% of all opioids prescribed to children 20 and under covered by Medicaid were dental-related. This is in line with existing research that for many children, dental related opioid prescriptions may be there first exposure to opioids.^10-12^ This early exposure may make those children more likely to misuse opioids throughout their lives.

The control of acute dental pain continues to contribute to the opioid prescription rate. In a systematic review, Moore and colleagues established that pain associated with third molar extraction is best controlled using a combination of acetaminophen and ibuprofen rather than with an opioid analgesic.^29^ Opioid analgesics are often not the most effective tool for pain relief and should not be considered a first-line choice. However, research shows this is still not the standard. For instance, between 2000 and 2010, 42% of Medicaid dental patients filled an opioid prescription within seven days of having a third molar tooth extraction.^30^ In another study, 65% of all dental-related opioids between 2013 and 2018 were prescribed for tooth extractions in a combined Medicaid/Commercial population, with third molars accounting for 40% of these extractions.^31^

While the CDC has not released specific opioid prescription guidelines for dental-related pain or even acute pain for that matter, it has released guidelines for chronic pain.^15^ These guidelines have been utilized by Suda and colleagues to evaluate opioid prescription patterns in commercial patients.^16^ 16% of dental-related Medicaid opioid prescriptions exceeded MMEs recommended for chronic pain in 2019. Regarding opioids that exceeded their recommended days supply, 42% were prescribed for over 3 days in 2019 for Medicaid patients. The results demonstrate that overprescribing has been on a consistent downward trend in Medicaid dentistry.

While much attention has been paid to opioid reduction strategies, little has been paid to their overall effectiveness. Since we do not know which states comprise our national sample, we are not able to evaluate specific strategies. Therefore, we cannot say to which degree reductions in both MME and the overall number of prescriptions is due to changes in provider behavior and education or rather state-led reduction programs. In a survey of dentists, a positive relationship was found between the prescribing culture of their residency program, and the prescribing patterns of the provider. Dentists who received instruction on opioid risks during residency were less likely to prescribe opioids to their patients.^32^

Research shows that states utilizing more robust prescription-drug monitoring programs (PDMPs) have fewer overall opioid overdose deaths than states whose programs are less robust.^33^ Our dataset does not identify the individual state, so it is impossible to link our results to specific PDMP regulations; however, their use is likely impacting these reductions.

## Limitations

Due to the structure of the dataset, we cannot directly link dental procedures to opioid prescriptions. Thus, we may be underestimating the true proportion of dental visits with an opioid prescription. In particular, though our results show the percentage of opioid prescriptions that were associated with a dental visit between 2012 and 2019 is 45.4%, the true number could be as high as 7.9%. This is due to the exclusion of some prescriptions from the final analysis as it was unclear whether a visit was dental- or medical-related. That is, it was unknown whether the prescriber was a medical professional or a dental professional, and so, as explained in the methods section, all prescriptions with a concurrent medical visit within three days were excluded. Furthermore, if a dentist prescribed an opioid, it was assumed filled within three days of the visit, consistent with other methods.^27^ Prescriptions filled outside that window were excluded from the analysis. Additionally, opioid claims were restricted to initial fills only and did not account for refill or repeat prescriptions.

Because this analysis uses claims data, it does not include procedures or prescriptions that were not paid or otherwise reflected in claims data. In addition, even though we excluded those dual eligibilities for Medicaid and Medicare, we may be missing some procedures or prescriptions that were paid for by an alternative insurance, as Medicaid is a payer of last resort. Due to limitations in the data, we are unable to identify the state policy environment in which the prescription was written.This limits our ability to identify the reasons for the decline in opioid prescriptions for dental procedures identified in our analysis or assess the impact of the particular policy environment in which opioids were prescribed.

## Conclusions

Our results are among the first to evaluate opioid prescribing patters for dental patients in a nationally-representative Medicaid population. They show a significant reduction in overall prescription rates, especially among non-elderly adults. The majority of opioid prescriptions are affiliated with surgical visits, with the percentage written for nonsurgical visits declining for each year in the study. The percent of prescriptions equal to or exceeding CDC guidelines of 50 MME/day declined across the study period. Although these decreases in both quantity and strength signify a positive trend, our analysis suggests that the overall rate is still too high and prescriptions are being written unnecessarily when an alternative treatment would be more effective in managing pain and reducing the incidence of opioid use disorder.

## Data Availability

The data is available through IBM Watson through a data use agreement.

## Acknowledgements

We would like to sincerely thank Avery Brow his contributions in writing assistance, editing and proofreading of this publication.

## Appendix A.

**Opioid prescription rates that exceed the CDC’s recommendations for chronic pain**. The [CDC](https://www.cdc.gov/mmwr/volumes/65/rr/rr6501e1.htm?CDC_AA_refVal=https%3A%2F%2Fwww.cdc.gov%2Fmmwr%2Fvolumes%2F65%2Frr%2Frr6501e1er.htm) recommends prescribing no more than 120 morphine milligram equivalent (MME) units per prescription as well as a maximum of 3 days’ supply for chronic pain.

| Year | Total Prescriptions | Over 120 Total MME | % over 120 MME | Over 3 Day Supply | % Over 3 Day Supply |
| --- | --- | --- | --- | --- | --- |
| 2012 | 213,710 | 70,790 | 33.1% | 96,091 | 45.0% |
| 2013 | 210,762 | 63,846 | 30.3% | 96,407 | 45.7% |
| 2014 | 246,721 | 70,701 | 28.7% | 111,722 | 45.3% |
| 2015 | 269,533 | 54,324 | 20.2% | 95,984 | 35.6% |
| 2016 | 242,536 | 63,799 | 26.3% | 103,118 | 42.5% |
| 2017 | 203,295 | 49,785 | 24.5% | 84,927 | 41.8% |
| 2018 | 139,567 | 26,710 | 19.1% | 57,612 | 41.3% |
| 2019 | 110,861 | 18,201 | 16.4% | 46,126 | 41.6% |
| Total | 1,636,985 | 418,156 | 25.5% | 691,987 | 42.3% |

## Appendix B.

Dental opioid prescription counts and respective percentages as defined by their active ingredient.

Appendix B. Dental opioid prescription counts and respective percentages as defined by their active ingredient .
| Prescription Name, Generic | 2012 | 2013 | 2014 | 2015 | 2016 | 2017 | 2018 | 2019 |
| --- | --- | --- | --- | --- | --- | --- | --- | --- |
| **Acetaminophen/Hydrocodone Bitartrate** | 194,713<br>(63.6%) | 188,424<br>(63.0%) | 205,571<br>(62.2%) | 213,212<br>(60.2%) | 193,169<br>(59.8%) | 165,849<br>(60.0%) | 113,066<br>(60.0%) | 93,076<br>(60.2%) |
| **Acetaminophen/Codeine Phosphate** | 50,108<br>(16.4%) | 47,606<br>(15.9%) | 50,616<br>(15.3%) | 57,472<br>(16.2%) | 55,033<br>(17.0%) | 49,053<br>(17.8%) | 36,582<br>(19.4%) | 32,733<br>(21.2%) |
| **Acetaminophen/Oxycodone Hydrochloride** | 44,199<br>(14.4%) | 44,879<br>(15.0%) | 53,318<br>(16.1%) | 61,911<br>(17.5%) | 54,199<br>(16.8%) | 42,821<br>(15.5%) | 24,950<br>(13.2%) | 17,768<br>(11.5%) |
| **Tramadol Hydrochloride** | 10,138<br>(3.3%) | 10,823<br>(3.6%) | 12,765<br>(3.9%) | 12,904<br>(3.6%) | 13,006<br>(4.0%) | 11,874<br>(4.3%) | 8,476<br>(4.5%) | 5,900<br>(3.8%) |
| **Oxycodone Hydrochloride** | 1,813<br>(0.6%) | 2,248<br>(0.8%) | 2,876<br>(0.9%) | 3,289<br>(0.9%) | 3,522<br>(1.1%) | 3,359<br>(1.2%) | 2,972<br>(1.6%) | 3,483<br>(2.3%) |
| **Hydrocodone Bitartrate/Ibuprofen** | 2,371<br>(0.8%) | 2,502<br>(0.8%) | 2,601<br>(0.8%) | 2,398<br>(0.7%) | 1,760<br>(0.5%) | 1,252<br>(0.5%) | 821<br>(0.4%) | 512<br>(0.3%) |
| **Acetaminophen/Tramadol Hydrochloride** | 573<br>(0.2%) | 652<br>(0.2%) | 827<br>(0.3%) | 1076<br>(0.3%) | 746<br>(0.2%) | 829<br>(0.3%) | 740<br>(0.4%) | 505<br>(0.3%) |
| **Morphine Sulfate** | 557<br>(0.2%) | 498<br>(0.2%) | 551<br>(0.2%) | 566<br>(0.2%) | 540<br>(0.2%) | 453<br>(0.2%) | 368<br>(0.2%) | 286<br>(0.2%) |
| **Fentanyl** | 694<br>(0.2%) | 575<br>(0.2%) | 542<br>(0.2%) | 430<br>(0.1%) | 228<br>(0.1%) | 168<br>(0.1%) | 24<br>(0.0%) | 13<br>(0.0%) |
| **Meperidine Hydrochloride** | 377<br>(0.1%) | 322<br>(0.1%) | 303<br>(0.1%) | 313<br>(0.1%) | 295<br>(0.1%) | 229<br>(0.1%) | 179<br>(0.1%) | 113<br>(0.1%) |

## Appendix C.

**Codeine and Tramadol prescription rates in children under 12 for the Dental Medicaid population**. The [FDA](https://www.fda.gov/drugs/drug-safety-and-availability/fda-drug-safety-communication-fda-restricts-use-prescription-codeine-pain-and-cough-medicines-and) released guidelines in Q2 of 2017 recommending against prescribing Codeine or Tramadol to children under 12 for pain management. These medicines are associated with difficulty breathing and other potentially life-threatening complications, with effects most pronounced in children under 12.

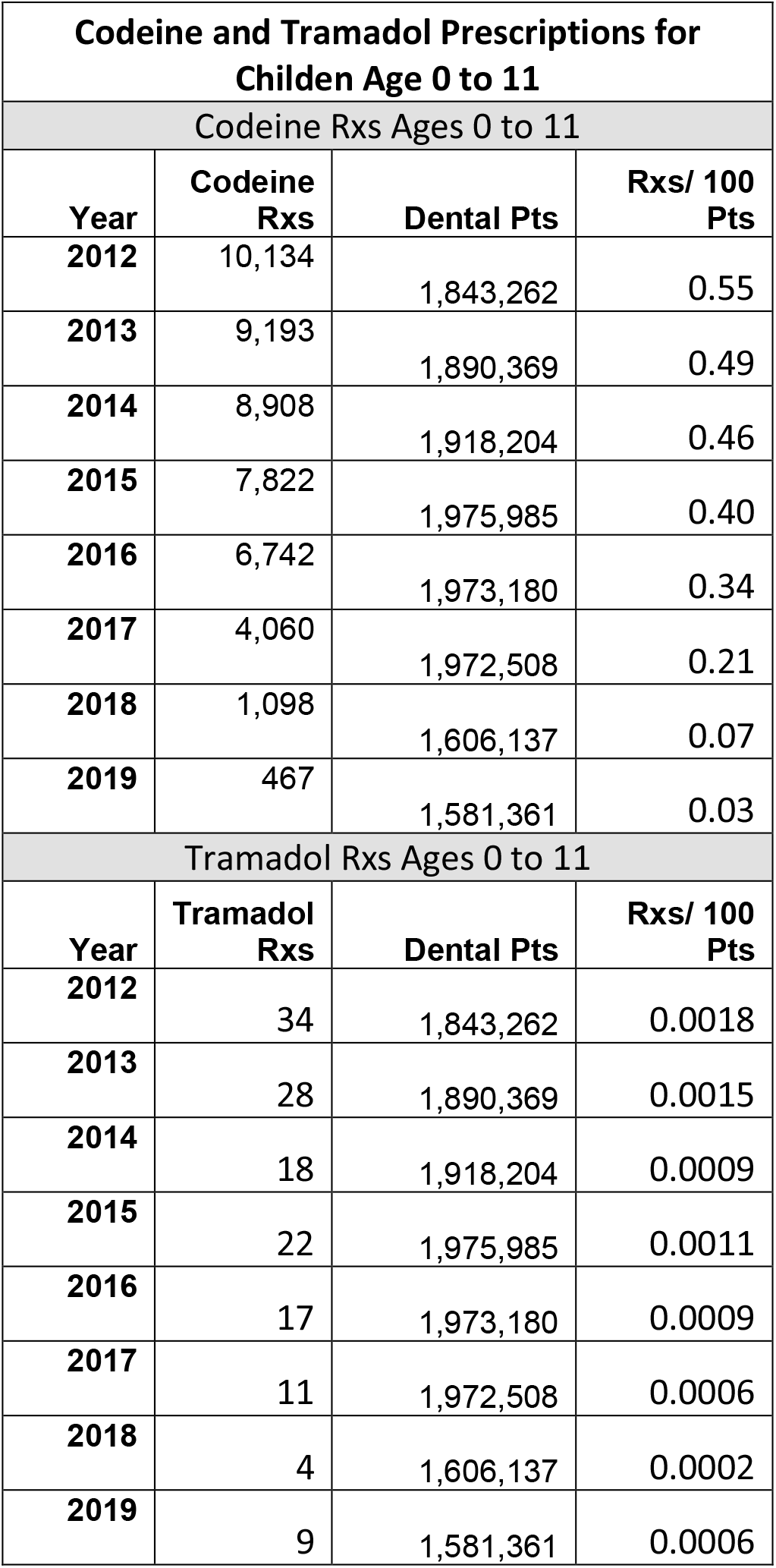

## References

1. Centers for Disease Control and Prevention. Opioid Overdose: U.S. Prescribing Rates Maps. CDC; 2018. https://www.cdc.gov/drugoverdose/maps/rxrate-maps.html Accessed Jul. 15.2020.

2. Centers for Disease Control and Prevention. Opioid Overdose. CDC; 2019. “https://www.cdc.gov/drugoverdose/index.html” Accessed Jul. 15, 2020.

3. Center BP. Tracking Federal Funding to combat the opioid crisis. Washington (DC): Bipartisan Policy Center. 2019 Mar. https://bipartisanpolicy.org/wp-content/uploads/2019/03/Tracking-Federal-Funding-to-Combat-the-Opioid-Crisis.pdf Accessed Jul. 15.2020.

4. Leslie DL, Ba DM, Agbese E, Xing X, Liu G. The economic burden of the opioid epidemic on states: the case of Medicaid. Am J Manag Care. 2019 Jul 1;25(13 Suppl):S243–9.

5. Davenport S, Weaver A, Caverly M. Economic impact of non-medical opioid use in the United States. SOA website. Published October 2019. https://www.soa.org/globalassets/assets/files/resources/research-report/2019/econ-impact-non-medical-opioid-use.pdf. Accessed March 5, 2021.

6. Volkow ND, McLellan TA, Cotto JH, Karithanom M, Weiss SB. Characteristics of opioid prescriptions in 2009. JAMA 2011;305(13):1299–301

7. Janakiram C, Chalmers NI, Fontelo P, et al. Sex and race or ethnicity disparities in opioid prescriptions for dental diagnoses among patients receiving Medicaid [published correction appears in J Am Dent Assoc. 2019 Oct;150(10):e135-e144]. J Am Dent Assoc. 2018;149(4):246–255.

8. Steinmetz CN, Zheng C, Okunseri E, Szabo A, Okunseri C. Opioid analgesic prescribing practices of dental professionals in the United States. JDR Clinical & Translational Research. 2017;2(3):241–8.

9. Nadeau R, Hasstedt K, Sunstrum AB, Wagner C, Tu H. Addressing the opioid epidemic: impact of opioid prescribing protocol at the University of Minnesota School of Dentistry. Craniomaxillofacial Trauma & Reconstruction 2018;11(2):104–10.

10. McCabe SE, West BT, Boyd CJ. Leftover prescription opioids and nonmedical use among high school seniors: a multi-cohort national study. Journal of Adolescent Health 2013;52(4):480–5.

11. McCauley JL, Hyer JM, Ramakrishnan VR, Leite R, Melvin CL, Fillingim RB, Frick C, Brady KT. Dental opioid prescribing and multiple opioid prescriptions among dental patients: administrative data from the South Carolina prescription drug monitoring program. JADA 2016;147(7):537–44.

12. Miech R, Johnston L, O’Malley PM, Keyes KM, Heard K. Prescription opioids in adolescence and future opioid misuse. Pediatrics 2015;136(5):e1169–77.

13. Maughan BC, Hersh EV, Shofer FS, Wanner KJ, Archer E, Carrasco LR, Rhodes KV. Unused opioid analgesics and drug disposal following outpatient dental surgery: a randomized controlled trial. Drug and alcohol dependence 2016;168:328–34.

14. Dionne RA, Gordon SM, Moore PA. Prescribing Opioid Analgesics for Acute Dental Pain: Time to Change Clinical Practices in Response to Evidence and Misperceptions. Compendium of continuing education in dentistry (Jamesburg, NJ: 1995) 2016;37(6):372–8.

15. Dowell D, Haegerich TM, Chou R. CDC Guideline for Prescribing Opioids for Chronic Pain – United States, 2016. Centers for Disease Control and Prevention;2016. “https://www.cdc.gov/mmwr/volumes/65/rr/rr6501e1.htm” Accessed Jul. 15, 2020.

16. Suda KJ, Zhou J, Rowan SA, McGregor JC, Perez RI, Evans CT, Gellad WF, Calip GS..Overprescribing of Opioids to Adults by Dentists in the U.S., 2011–2015. American Journal of Preventive Medicine 2020.

17. Lev R, Lee O, Petro S, Lucas J, Castillo EM, Vilke GM, Coyne CJ. Who is prescribing controlled medications to patients who die of prescription drug abuse?. The American journal of emergency medicine 2016;34(1):30–5.

18. De Rossi SS. Dentistry and the opioid epidemic: do dentists over prescribe opioids? Conscious Sedation Consulting; 2016. https://www.sedationconsulting.com/3350-2/ Accessed Jul. 15, 2020.

19. Chung CP, Callahan ST, Cooper WO, Dupont WD, Murray KT, Franklin AD, Hall K, Dudley JA, Stein CM, Ray WA. Individual short-acting opioids and the risk of opioid-related adverse events in adolescents. Pharmacoepidemiology and drug safety 2019;28(11):1448–56.

20. U.S. Food and Drug Administration. FDA Drug Safety Communication: FDA restricts use of prescription codeine pain and cough medicines and tramadol pain medicines in children; recommends against use in breastfeeding women. Silver Spring, MD: FDA; 2018. “https://www.fda.gov/drugs/drug-safety-and-availability/fda-drug-safety-communication-fda-restricts-use-prescription-codeine-pain-and-cough-medicines-and” Accessed Jul. 15, 2020.

21. Obadan-Udoh E, Lupulescu-Mann N, Charlesworth CJ, Muench U, Jura M, Kim H, Schwarz E, Mertz E, Sun BC. Opioid prescribing patterns after dental visits among beneficiaries of Medicaid in Washington state in 2014 and 2015. JADA 2019;150(4):259–68.

22. Ali MM, Cutler E, Mutter R, Henke RM, O’Brien PL, Pines JM, Mazer-Amirshahi M, Diou-Cass J. Opioid use disorder and prescribed opioid regimens: evidence from commercial and Medicaid claims, 2005–2015. Journal of medical toxicology 2019;15(3):156–68.

23. Fernandes JC, Campana D, Harwell TS, Helgerson SD. High mortality rate of unintentional poisoning due to prescription opioids in adults enrolled in Medicaid compared to those not enrolled in Medicaid in Montana. Drug and alcohol dependence 2015;153:346–9.

24. IBM. IBM MarketScan Research Databases.”https://www.ibm.com/us-en/marketplace/marketscan-research-databases?lnk=hm)” Accessed Jul. 15, 2020.

25. Roberts SM, Wilson CF, Seale NS, McWhorter AG. Evaluation of morphine as compared to meperidine when administered to the moderately anxious pediatric dental patient. Pediatr Dent. 1992;14(5):306–313.

26. NIDA. 2020, June 3. Opioid Addiction. Retrieved from https://www.drugabuse.gov/publications/principles-drug-addiction-treatment-research-based-guide-third-edition/evidence-based-approaches-to-drug-addiction-treatment/pharmacotherapies/opioid Accessed Jul. 15, 2020.

27. Gupta N, Vujicic M, Blatz A. Opioid prescribing practices from 2010 through 2015 among dentists in the United States: what do claims data tell us?. JADA 2018;149(4):237–45.

28. Centers for Disease Control and Prevention. Calculating total daily dose of opioids for safer dosage. “https://www.cdc.gov/drugoverdose/resources/data.html” Accessed Mar. 2, 2021.

29. Moore PA, Hersh EV. Combining ibuprofen and acetaminophen for acute pain management after third-molar extractions: translating clinical research to dental practice. JADA 2013;144(8):898–908.

30. Baker JA, Avorn J, Levin R, Bateman BT. Opioid prescribing after surgical extraction of teeth in Medicaid patients, 2000-2010. Jama 2016;315(15):1653–4.

31. Chua KP, Hou HM, Waljee JF, Brummett CM, Nalliah RP. Opioid prescribing patterns by dental procedure among US publicly and privately insured patients, 2013 through 2018. JADA 2021.

32. McCauley JL, Leite RS, Melvin CS, Fillingam RB, Brady KT. Dental opioid prescribing practices and risk mitigation strategy implementation: identification of potential targets for provider-level intervention. Substance Abuse 2016;37(1):9–14.

33. Pardo B. Do more robust prescription drug monitoring programs reduce prescription opioid overdose?. Addiction 2017;112(10):1773–83.

